# Community Antibiotic Prescribing in Patients with COVID-19 Across Three Pandemic Waves: A Population-Based Cohort Study

**DOI:** 10.1101/2023.06.29.23291797

**Authors:** Laura Ciaccio, Peter T Donnan, Benjamin J Parcell, Charis A Marwick

**Author notes:** **<u>Corresponding Author</u>:** Laura Ciaccio, University of Dundee, School of Medicine, Mailbox 1, Level 7, Corridor L, Ninewells Hospital, Dundee, DD1 9SY.

## Abstract

**Background:** Reported changes in antibiotic prescribing during the COVID-19 pandemic have focused on hospital prescribing or community population trends. Community antibiotic prescribing for individuals with COVID-19 are less well described.

**Methods:** Data covering a complete geographic population (∼800,000) were utilised. SARS-CoV-2 virus test results from February 1, 2020-March 31, 2022 were included. Anonymised data were linked to prescription data +/-28 days of the test, GP data for high-risk comorbidities, and demographic data. Multivariate binary logistic regression examined associations between patient factors and the odds of antibiotic prescription.

**Results:** Data included 768,206 tests for 184,954 individuals, identifying 16,240 COVID-19 episodes involving 16,025 individuals. There were 3,263 antibiotic prescriptions +/-28 days for 2,385 patients. 35.6% of patients had a prescription only before the test date, 52.5% of patients after, and 11.9% before and after. Antibiotic prescribing reduced over time: 20.4% of episodes in wave one, 17.7% in wave two, and 12.0% in wave three. In multivariate logistic regression, being female (OR 1.31, 95% CI 1.19,1.45), older (OR 3.02, 95% CI 2.50, 3.68 75+ vs <25 years), having a high-risk comorbidity (OR 1.45, 95% CI 1.31, 1.61), a hospital admission +/-28 days of an episode (OR 1.58, 95% CI 1.42, 1.77), and health board region (OR 1.14, 95% CI 1.03, 1.25, board B versus A) increased the odds of receiving an antibiotic.

**Conclusion:** Community antibiotic prescriptions in COVID-19 episodes were uncommon in this population and likelihood was associated with patient factors. The reduction over pandemic waves may represent increased knowledge regarding COVID-19 treatment and/or evolving symptomatology.

## Introduction

Antibiotic surveillance and stewardship remain priorities during viral pandemics,^1^ with the majority of antibiotics prescribed in the community.

Many studies on antibiotic use during the COVID-19 pandemic have focused on hospitalised patients. Systematic reviews report high rates of antibiotic prescriptions early in the pandemic at around 70%, despite bacterial co-infection being confirmed in less than 10% of patients.^1–4^

Studies of community antibiotic prescribing have largely described overall changes at the population level, particularly during the first pandemic wave.^5–9^ NHS England reported antibiotic prescriptions decreased 15.5% from April 1 to August 31, 2020, compared to the same period in 2019 but, adjusted for the reduction in appointments, this represented an increase of 6.7%.^9^ At the local start of the pandemic in March 2020, Scotland saw a 44% increase in community prescriptions for antibiotics commonly used to treat respiratory infections compared to 2019, but this dropped to 34% below the 2019 rate by May 2020.^10^

Community antibiotic prescribing for individuals with COVID-19 is less well studied.^4,10,11^ Of two previous relevant studies, in the US^11^ and Italy,^12^ one used diagnostic codes rather than cases confirmed by testing and was limited to patients using one medical insurance provider.^11^ Both examined short time intervals before and after the diagnosis (so may have under-estimated prescribing), and both covered only the first two pandemic waves.

This study aimed to examine community antibiotic prescribing rates across a complete geographic area for people with a positive COVID-19 test across three pandemic waves, and to examine health and demographic factors associated with antibiotic prescribing.

## Methods

Anonymised data from two National Health Service Scotland (NHS) Health Boards covering approximately 20% of the Scottish population (n= 863,974) were accessed via a University of Dundee Health Informatics Centre secure remote desktop. Datasets were linked at the individual level using the Community Health Index (CHI) number, a unique identifier used to identify patients across all NHS Scotland healthcare episodes.

COVID-19 test results from 28^th^ February 2020 (date of the first COVID-19 positive test in Scotland) ^13^ to 31^st^ March 2022 included Polymerase Chain Reaction (PCR) test and Lateral Flow Test (LFT) results from NHS and private (with NHS contracts) laboratories and at-home tests centrally reported. Multiple tests per patient on the same day were deduplicated, and repeated positive results within 90 days were considered the same episode of COVID-19, in accordance with NHS Scotland testing guidance.^14^

All COVID-19 episodes were linked to community antibiotic prescriptions, demography, high-risk comorbidity/shielding, and hospital admission data. The community prescribing dataset captures dispensed prescribed items (“prescriptions”) using pharmacy claims for reimbursement. Prescriptions for all oral antibiotics in the British National Formulary (BNF), Chapter 5, subsections 5.1-Antibacterial Drugs, were included. Prescriptions from 28 days prior (−28 to -1 days) to 28 days post-(0 to +28 days) positive test were included to capture pharmacy claims data batched monthly.

Demography data included calculated age, sex, health board, and Scottish Index of Multiple Deprivation quintile (SIMD5), which is based on residential postcode. Quintile 1 is the most deprived and 5 the least.

The high-risk comorbidity dataset included patients flagged in primary care records for possible shielding advice based on diagnoses and/or prescriptions. The conditions included asthma, chronic obstructive pulmonary disorder (COPD), diabetes, hypertension, ischemic heart disease, other respiratory conditions, and immunological conditions (supplementary Table S1). All patients flagged for these conditions were considered to have high-risk comorbidity.

Hospital admissions were extracted from the Scottish Morbidity Record 01 (SMR01) dataset. Admissions with a discharge date from 28 days prior to 28 days post-positive test date were included.

COVID-19 episodes were categorised into pandemic waves as previously defined for Scotland,^15^ with the end dates of each wave extended to prevent gaps in the study period. Wave 1 started 28th February 2020, wave 2 1st August 2020, and wave 3 1st May 2021. Individuals could have more than one episode, in one or more waves.

Univariate and multivariate binary logistic regression analyses were used to examine associations between health and demographic factors and the likelihood of receiving a community antibiotic prescription for that COVID-19 episode. Variables included age (<25, 25-44, 45-64, 65-74, 75+ years), gender, health board of residence (A vs B), high-risk comorbidity, hospital admissions +/-28 days, Scottish Index of Multiple Deprivation (SIMD5) quintile, and pandemic wave. All variables were included in multivariate analysis due to social and/or clinical relevance.

All analyses used RStudio version 4.1.2.

## Results

The dataset included 768,206 tests for 184,954 individuals (21.4% of the population). There were 16,240 COVID-19 episodes involving 16,025 individuals. 98.7% of included individuals had one episode, 1.3% had two, and 0.01% had three. The mean age at episode was 51.9 years (SD 24.8), 59.4% were female, and 57.2% were in health board B (Table 1). The age distribution of episodes varied across waves, with 29.0% in wave 1 involving people aged 75+ years, compared to 18.2% in wave 3, and 3.0% in wave 1 aged <25 years compared to 23.0% in wave 3.

**Table 1:** Study demographics.

| Variable categories | | Total study population | Pts with $\geq 1$ test | Pts. with $\geq 1$ episode | COVID Episodes | | |
| --- | --- | --- | --- | --- | --- | --- | --- |
|  |  |  |  |  | Wave 1* | Wave 2* | Wave 3* |
| Total |  | 863974<br>(100%) | 184953<br>(21.4%) | 16025<br>(1.9%) | 2430<br>(0.3%) | 4324<br>(0.5%) | 9434<br>(1.1%) |
| Age group | <25 | 200441<br>(23.2%) | 30220<br>(16.3%) | 2632<br>(16.4%) | 81<br>(3.0%) | 385<br>(8.9%) | 2172<br>(23.0%) |
|  | 25-44 | 220996<br>(25.6%) | 41959<br>(22.7%) | 3888<br>(24.3%) | 537<br>(22.1%) | 940<br>(21.7%) | 2464<br>(26.1%) |
|  | 45-64 | 237115<br>(27.4%) | 52017<br>(28.1%) | 4337<br>(27.1%) | 849<br>(34.9%) | 1222<br>(28.3%) | 2313<br>(24.5%) |
|  | 65-74 | 101797<br>(11.8%) | 23783<br>(12.9%) | 1417<br>(8.8%) | 258<br>(10.6%) | 427<br>(9.9%) | 773<br>(8.2%) |
|  | 75+ | 103625<br>(12.0%) | 36974<br>(20.0%) | 3751<br>(23.4%) | 705<br>(29.0%) | 1350<br>(31.2%) | 1712<br>(18.2%) |
| Sex | Male | 426794<br>(49.4%) | 75605<br>(40.9%) | 6533<br>(40.8%) | 897<br>(36.9%) | 1549<br>(35.8%) | 4135<br>(43.8%) |
|  | Female | 437180<br>(50.6%) | 109348<br>(59.1%) | 9492<br>(59.2%) | 1533<br>(63.1%) | 2775<br>(64.2%) | 5299<br>(56.2%) |
| Any high-risk comorbidity | No | 630838<br>(73.0%) | 113573<br>(61.4%) | 9975<br>(62.2%) | 1454<br>(59.8%) | 2401<br>(55.5%) | 6221<br>(65.9%) |
|  | Yes | 233136<br>(27.0%) | 71381<br>(38.6%) | 6050<br>(37.8%) | 976<br>(40.2%) | 1923<br>(44.5%) | 3213<br>(34.1%) |
| Health board | A | 404094<br>(46.8%) | 67887<br>(36.7%) | 6882<br>(42.9%) | 779<br>(32.1%) | 1667<br>(38.6%) | 4473<br>(47.4%) |
|  | B | 459880<br>(53.2%) | 117066<br>(63.3%) | 9143<br>(57.1%) | 1651<br>(67.9%) | 2657<br>(61.4%) | 4961<br>(52.6%) |
| Scottish Index of Multiple Deprivation (SIMD) quintile | 1 (most deprived) | 135851<br>(15.7%) | 30411<br>(16.4%) | 3194<br>(19.9%) | 426<br>(17.5%) | 888<br>(20.5%) | 1922<br>(20.4%) |
|  | 2 | 145538<br>(16.8%) | 31357<br>(17.0%) | 2839<br>(17.7%) | 374<br>(15.4%) | 878<br>(20.3%) | 1604<br>(17.0%) |
|  | 3 | 153304<br>(17.7%) | 32195<br>(17.4%) | 2808<br>(17.5%) | 447<br>(18.4%) | 746<br>(17.3%) | 1650<br>(17.5%) |
|  | 4 | 203258<br>(23.5%) | 46426<br>(25.1%) | 3556<br>(22.2%) | 649<br>(26.7%) | 934<br>(21.6%) | 2007<br>(21.3%) |
|  | 5 (least deprived) | 152013<br>(17.6%) | 29995<br>(16.2%) | 2312<br>(14.4%) | 317<br>(13.0%) | 599<br>(13.9%) | 1417<br>(15.0%) |
|  | Missing Data | 74010<br>(8.6%) | 14569<br>(7.8%) | 1316<br>(8.2%) | 217<br>(8.9%) | 279<br>(6.5%) | 834<br>(8.8%) |

\*Individuals may appear in multiple waves and may have multiple episodes in a single wave if they occur >90 days apart

The most common comorbidities were hypertension, other respiratory disorders, and asthma. 17.4% of those with at least one episode had one high-risk comorbidity, and 2.3% of patients had 4 or more (Figure 1).

**Figure 1.**
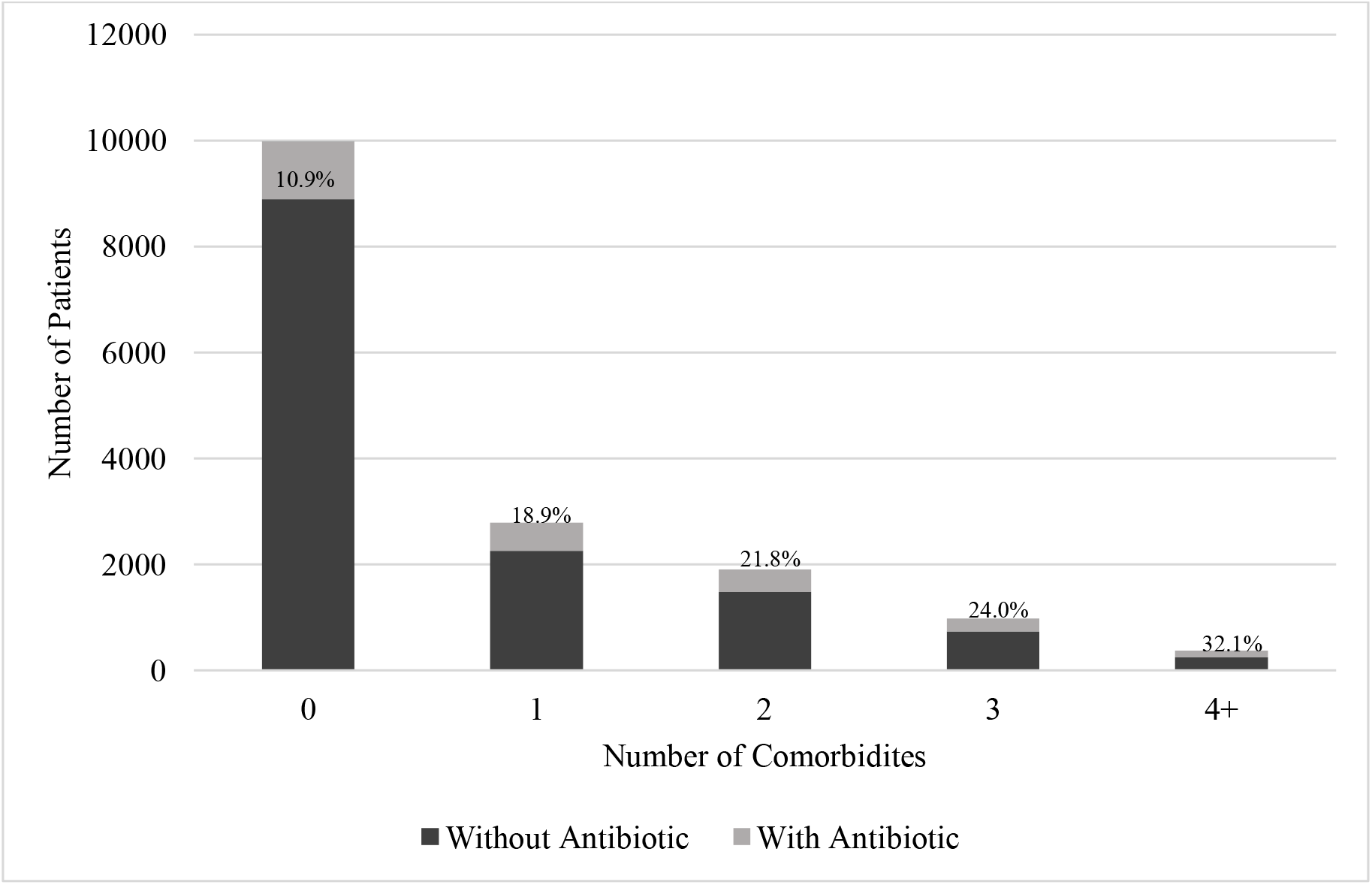
Distribution of high-risk comorbidities for individuals with a COVID-19 episode and proportion (%) with an antibiotic prescription in each category.

**Figure 2.**
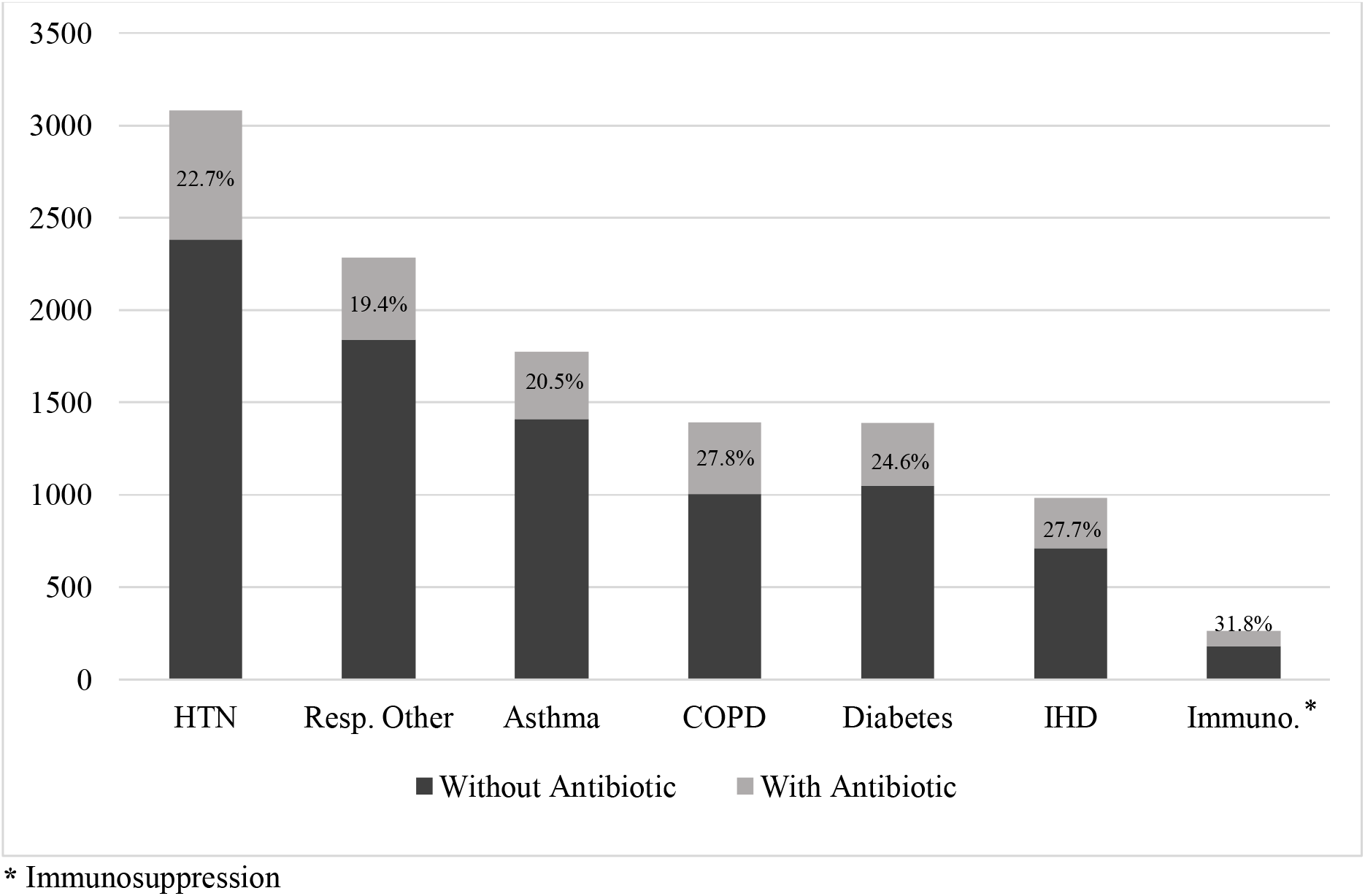
Antibiotic prescriptions for individuals with a COVID-19 episode by high-risk comorbidity (%)

There were 3,263 antibiotic prescriptions within 28 days of 2,395 (18.1%) episodes. 853 (35.6%) episodes had an antibiotic prescription before the test only, 1,252 (52.3%) after only, and 290 (12.1%) both before and after. The number of antibiotic prescriptions per episode ranged from 1 to 8, but the majority (54.5%) had one.

Antibiotic prescribing in COVID-19 reduced over time, at 20.4% of episodes in wave one, 17.7% in wave two, and 12.0% in wave three (Table 2). Amoxicillin (22.5%) and doxycycline (15.1%) were most prescribed overall, accounting for 41.1% of antibiotic prescriptions in wave 1, 34.8% in wave 2, and 37.8% in wave 3 (Table 1).

**Table 2:** Testing and Antibiotic Frequencies by UK COVID-19 Wave.

|  | Total tests | Mean tests per month | Total episodes | Episodes with antibiotic prescription(s) +/-28 days | Proportion of episodes with an antibiotic prescription +/-28 day |
| --- | --- | --- | --- | --- | --- |
| Wave 1 | 47606 | 3967 | 2432 | 496 | 20.4% |
| Wave 2 | 432278 | 48031 | 4330 | 765 | 17.7% |
| Wave 3 | 288232 | 24019 | 9478 | 1134 | 12.0% |
| Total | 768206 | 30728 | 16240 | 2395 | 14.7% |

**Wave 1:** February 28, 2020- July 31, 2020; **Wave 2:** August 1, 2020- April 30, 2021; **Wave 3:** May 1, 2021-March 31, 2022 (end of study period)

In univariate logistic regression, all variables were significantly associated with the odds of having a prescription (Table 3). In multivariate analysis, being female (OR 1.31, 95%CI 1.19 to 1.45), older (OR 3.02 [2.50 to 3.68] for 75+ vs <25 years), having a high-risk comorbidity (Figure 1) (OR 1.45 [1.31 to 1.61]), having a hospital admission within 28 days of an episode (OR 1.58 [1.42 to 1.77]), and living in health board B rather than A (OR 1.14 [1.03 to 1.25]) significantly increased the likelihood of receiving an antibiotic (Table 3). Having an episode in wave 2 (OR 0.86, 95% CI 0.75 to 0.99) or wave 3 (OR 0.71, 95% CI 0.62 to 0.81) significantly decreased the odds of receiving an antibiotic prescription compared to wave 1. Associations with deprivation did not show a clear trend (Table 3).

**Table 3:** Associations between demographic and healthcare factors and the odds of a community antibiotic prescription for COVID-19 episodes, from binary logistic regression.

| Variable | Category | Frequency | Univariate OR (95% CI) | p-value | Multivariate OR (95% CI) | p-value |
| --- | --- | --- | --- | --- | --- | --- |
| Age Group | <25 | 2652 | REF | - | - | - |
|  | 25-44 | 3958 | 1.53 (1.28, 1.84) | <0.001 | 1.35 (1.11, 1.64) | 0.003 |
|  | 45-64 | 4394 | 1.88 (1.58, 2.24) | <0.001 | 1.51 (1.25, 1.84) | <0.001 |
|  | 65-74 | 1431 | 2.83 (2.32, 3.48) | <0.001 | 1.91 (1.52, 2.39) | <0.001 |
|  | 75+ | 3805 | 4.89 (4.15, 5.78) | <0.001 | 3.02 (2.50, 3.68) | <0.001 |
| Sex | Male | 6599 | REF | - | - | - |
|  | Female | 9641 | 1.25 (1.15, 1.37) | <0.001 | 1.31 (1.19, 1.45) | <0.001 |
| High-Risk Comorbidity | No | 10108 | REF | - | - | - |
|  | Yes | 6132 | 2.34 (2.05, 2.44) | <0.001 | 1.45 (1.31, 1.61) | <0.001 |
| Hospital Admission +/-28 days | No | 13047 | REF | - | - | - |
|  | Yes | 3193 | 2.28 (2.07, 2.51) | <0.001 | 1.58 (1.42, 1.77) | <0.001 |
| Health Board | A | 6943 | REF | - | - | - |
|  | B | 9297 | 1.15 (1.06, 1.26) | <0.001 | 1.14 (1.03, 1.25) | 0.01 |
| Scottish Index of Multiple Deprivation (SIMD) quintile | 1 (most deprived) | 3251 | REF | - | - | - |
|  | 2 | 2864 | 0.83 (0.72, 0.96) | 0.01 | 0.81 (0.70, 0.94) | 0.01 |
|  | 3 | 2849 | 1.07 (0.94, 1.24) | 0.29 | 1.00 (0.86, 1.15) | 0.99 |
|  | 4 | 3604 | 1.07 (0.94, 1.22) | 0.31 | 0.88 (0.77, 1.01) | 0.07 |
|  | 5 (least deprived) | 2341 | 0.93 (0.80, 1.09) | 0.38 | 0.84 (0.72, 0.98) | 0.03 |
| COVID Wave | 1 | 2432 | REF | - | - | - |
|  | 2 | 4330 | 0.84 (0.74, 0.95) | 0.005 | 0.86 (0.75, 0.99) | 0.03 |
|  | 3 | 9478 | 0.53 (0.47, 0.60) | <0.001 | 0.71 (0.62, 0.81) | <0.001 |

## Discussion

### Principal findings

In this large, population-based cohort study, we saw a changing patient profile for COVID-19 patients over time. We found a relatively low rate of community antibiotic prescriptions for COVID-19 episodes at 14.7%, with a reduction over time, from 20.4% of episodes in wave 1 to 12.0% in wave 3. We also found clear associations between individual demographic and healthcare factors and receipt of an antibiotic.

The decrease in antibiotic prescribing over time will have multiple contributing factors. Early COVID-19 treatment guidelines were modified during the pandemic as data regarding low levels of bacterial co-infection emerged.^16^ Testing patterns also changed, and, as testing became available to the public and mandated for many sectors, misdiagnosis as bacterial infections became less likely. Vaccination roll-out, with attenuated symptom severity, may have reduced presentations with COVID-19 to community medical services, public anxiety, and clinicians’ likelihood of prescribing an antibiotic. Emerging viral variants had different symptoms and/or severity,^17^ likely affecting antibiotic prescriptions similarly to vaccination. The high proportion of amoxicillin and doxycycline prescriptions aligns with Scottish guidance for the treatment of (presumed bacterial) respiratory tract infections.^18,19^ The proportion of prescriptions for these drugs was lowest in wave two, which may reflect the dominant variants in wave 2 (alpha and delta) having lower prevalence of respiratory symptoms.^17,20^

Older age was the strongest demographic predictor of antibiotic prescribing, likely due to higher testing rates and lower threshold for antibiotic prescriptions.^21^ Older patients are often less likely to be asymptomatic,^22^ and, in this study, had more hospital admissions and more comorbidity. Females were more likely to have a test, consistent with other studies,^23^ and had more positive tests and more antibiotic prescriptions. This may reflect differences in accessing medical care, with females reportedly contacting health services more often and earlier in an illness.^24^ The association between community prescribing and hospital admissions may reflect COVID-19 episodes requiring hospitalisation having longer symptoms and/or more concern and community healthcare contact. However, it may reflect more vulnerable individuals having more healthcare contact in general, rather than specific features of the COVID-19 episode.

### Comparison with other studies

There are very few studies examining community antibiotic prescribing in individuals with COVID-19, with more focused on changes in total community or hospital prescribing.^5–8,10^ Of 154 studies in a meta-analysis of antibiotic prescribing in COVID-19, 12 were mixed inpatient and outpatient settings, but none were community only.^4^ One outpatient study completed after the previous meta-analysis examined antibiotic prescriptions for American Medicare beneficiaries with prescription drug (Part D) coverage, with an outpatient visit, including the Emergency Department, from April 2020 to April 2021 with a primary diagnosis code of COVID-19 (U071). Of >1 million encounters, around 30% of patients received an antibiotic prescription within 7 days pre- or post-visit.^11^ This is higher than in our study (despite our longer time window pre/post-diagnosis), but the Medicare population was limited to those over 65 and to the first two pandemic waves, where we also observed higher rates. We have also been able to include all age groups, including children, rather than focusing on older individuals who were found to have greater odds of receiving an antibiotic prescription. The authors note their lack of data on underlying health conditions and hospital admissions as reported limitations,^11^ and we found these factors were influential in prescribing.

An Italian study examined community prescriptions for 331,704 individuals with laboratory-confirmed positive COVID-19 PCR tests from March 2020 to May 2021. Prescriptions were included from 3 days pre- to 7 days post-positive test. 23% of cases received an antibiotic, with a notable increase from 18% of cases in November 2020 to 31% in March 2021.^12^ The overall rate is higher than in our study, and the increase over time is contrary to our findings, but they did not include data from wave 3.

Studies examining trends in overall community antibiotic prescribing all report decreases across 2020. Quarterly US data reported an overall reduction, including a decrease of 44% in amoxicillin prescriptions, from calendar quarter two to quarter four.^25^ A study from Spain reported a decrease (pooled DDD reduction) in prescribed antibiotics of 7.6% in quarter one and 36.8% in quarter two of 2020 compared to the same time in 2019.^8^ Similarly, France and Canada reported overall reductions of 18.2% and 31.2%, respectively, in outpatient antibiotic prescriptions in 2020 compared to 2019.^7,26^

In hospital settings, early COVID-19 systematic reviews found that bacterial co-infections were confirmed for only around 7-8% of patients, but 70-72% received an antibiotic.^1–3^ An April 2020 survey in Scottish hospitals found that 38.3% of patients with suspected or confirmed COVID-19 were prescribed an antibiotic.^27^ These rates are higher than we observed, but the threshold for antibiotic prescribing will be lower in hospitalised patients, who are more unwell and higher risk, and they are from earlier in the pandemic.

### Strengths and limitations

Key strengths of this study are the size of the population-level dataset and the use of administrative data, which increases generalizability and minimises the impact of any missing data. SIMD was the only variable with notable missing data (8%). These data were missing completely at random, with affected individuals evenly distributed across categories of other variables, not affecting the findings or interpretation. Another key strength is the universal use of CHI numbers (de-identified) across all NHS services, enabling multiple datasets to be linked longitudinally. The timeframe of this study allows analysis of trends over multiple pandemic waves. A limitation of this study is that data on the coded indication for the antibiotic prescriptions were unavailable, so we have potentially included antibiotics prescribed for other conditions. However, primary care coding is of variable quality and utility for research,^28^ and patients presenting with a febrile illness coded as something else but subsequently diagnosed as COVID-19 would be missed by coding. Bacteriology data were not included, and some individuals may have had bacterial secondary or co-infection with appropriate antibiotic treatment. However, most patients with presumed bacterial respiratory tract infections, including pneumonia, never have a bacteriological diagnosis,^29^ and some bacterial pathogens can also be commensals, so bacteriology data are unlikely to facilitate evaluation of appropriateness at the population level. Data on individual COVID-19 vaccination status were not available, but vaccine uptake was high in the study population (>/=85% of eligible population had 2+ doses).^13^ The findings may not be generalisable to areas with different demographic characteristics (ethnicity data were unavailable), but the study regions are demographically representative of the Scottish population. Clinical outcomes of COVID-19 episodes were not included but would be prone to confounding by indication, as sicker patients would be more likely to both get an antibiotic prescription and have an adverse outcome.

### Implications for Policy and Practice

The decreasing use of antibiotics found in each subsequent COVID-19 wave suggests that prescribers and the public responded to changing guidance and recommendations and became better at recognising and managing COVID-19. Reducing unnecessary antibiotic prescriptions supports antimicrobial stewardship, and the findings align with previous work indicating that vaccines to reduce symptomatic illness, including viral, can reduce antibiotic prescribing.^30^ The difference in prescribing between health boards presents an opportunity for sharing good stewardship practice.

Although this study used rich, linked administrative data, GP consultation data are not routinely available in Scotland, and, despite the limitations, this gap should be addressed to support surveillance and research to inform practice, for example, on appropriateness of antibiotic prescriptions.

### Implications for Future Research

This work highlights the need for more research on community management of individuals with COVID-19, and the drivers of potentially unnecessary antibiotic prescribing. Qualitative work with prescribers in the community could enhance understanding of practice changes over time, and with individuals who had COVID-19 could enhance understanding of changes in healthcare-seeking behaviour or access. It would also be of interest to examine whether changes in community prescribing observed in COVID-19 are replicated for other viral illnesses. These findings could inform antimicrobial stewardship and strategies, including in future viral pandemics.

## Conclusions

Community antibiotic prescriptions in people with COVID-19 were relatively uncommon in this study population and were associated with increased age and comorbidity. There was a significant reduction over time which may represent increased knowledge and experience of COVID-19 and/or decreased symptom severity due to vaccination and changes in the dominant variants of the virus over time.

## Data Availability

Data from this study are not publically available but may be accessed by approved researchers via application to the Health Informatics Centre at the University of Dundee.

## Transparency declaration

The authors have no conflicts of interest to declare.

## Author contributions

Concept and design: All authors. Interpretation of data: All authors. Drafting of the manuscript: LC. Manuscript revision: All authors. Statistical analysis: LC. Administrative, technical, or material support: All authors.

## Acknowledgements

LC was funded by the Medical Research Foundation National PhD Training Programme in Antimicrobial Resistance Research (MRF-145-0004-TPG-AVISO)

