## Supplemental Table 1 for "Community Antibiotic Prescribing in Patients with COVID-19 Across Three Pandemic Waves: A Population-Based Cohort Study"

**Supplementary data**

**Table S1- Comorbidity categories used in study**

| Category in source data | Study comorbidity group | Count in overall population |
| --- | --- | --- |
| Diagnosis Asthma | Asthma | 66041 |
| Diagnosis COPD^1^ | COPD | 18018 |
| Diagnosis Cystic Fibrosis | Respiratory- other* | 69 |
| Diagnosis Diabetes | Diabetes | 40803 |
| Diagnosis Hypertension | Hypertension | 107696 |
| Diagnosis Ischemic Heart Disease | Ischemic Heart Disease | 27511 |
| Diagnosis Transplant | Immunosuppression | 479 |
| Diagnosis Lung Disease | Respiratory- other* | 97401 |
| Diagnosis Severe Lung Disease | Respiratory- other | 10098 |
| Indication Transplant | Immunosuppression | 1562 |
| Indication Immuno | Immunosuppression | 770 |
| Indication Respiratory | Respiratory- other* | 6103 |
| Asthma Prescriptions: HS ICS^2^, ICS LABA^3^, LABA^4^, Montel, Prednisone | Asthma | 30477 |
| COPD Prescription: ICS, LABA^3^, LAMA^5^, Roflumilast | COPD | 66517 |
| Immuno Cortisol 5 or 20 Prescription | Immunosuppression | 4365 |
| Transplant DMARD Prescription | Immunosuppression | 1562 |
| Azithromycin Prescription | Respiratory- other* | 1109 |
| Immuno DMARD^6^ Prescription | Immunosuppression | 4247 |

* only if patient was not found to also have asthma, COPD, or cystic fibrosis, ^1^ Chronic obstructive pulmonary disease, ^2^ High strength inhaled corticosteroid, ^3^ Inhaled corticosteroid long-acting bronchodilator, ^4^ Long-acting bronchodilator, ^5^ Long-acting muscarinic antagonist, ^6^ Disease modifying anti-rheumatic drugs
